# The Silent Author in Urology: Quantifying Large Language Model Influence at the Corpus Level

**DOI:** 10.1101/2025.07.21.25331681

**Authors:** Adel Arezki, James Man Git Tsui, Wassim Kassouf

## Abstract

**Introduction:** The recent increasing use of large language models (LLMs) such as ChatGPT in scientific writing raises concerns about authenticity and accuracy in biomedical literature. This study aims to quantify the prevalence of Artificial Intelligence (AI)-generated text in urology abstracts from 2010 to 2024 and assess its variation across journal categories and impact factor quartiles.

**Methods:** A retrospective analysis was conducted on 64,444 unique abstracts from 38 journals in the field of urology published between 2010-2024 and retrieved via the Entrez API from PubMed. Abstracts were then categorized by subspecialty and stratified by impact factor (IF). A synthetic reference corpus of 10,000 abstracts was generated using GPT-3.5-turbo. A mixture model estimated the proportion of AI-generated text 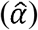 annually, using maximum likelihood estimation with Laplace smoothing. Calibration and specificity of the estimator were assessed using pre-LLM abstracts and controlled mixtures of real and synthetic text. Statistical analyses were performed using Python 3.13.

**Results:** The proportion of AI-like text was negligible from 2010 to 2019, rising to 1.8% in 2020 and 5.3% in 2024. In 2024, Men’s Health journals showed the highest AI-like text, while Oncology journals had the lowest. Journals in the highest and lowest IF quartiles showed a higher proportion of AI-like text than mid-quartiles. Type-Token Ratio (TTR) remained stable across the study period. Our validation calibration set showed an area under the curve of 0.5326.

**Conclusion:** Textual patterns similar to AI-generated language has risen sharply in urology abstracts since ChatGPT’s release in late 2022, and this trend varies by journal type and IF.

**Patient summary:** We looked at whether large language models (LLM) such as ChatGPT are influencing the way urology research is written. We found that signs of LLM-like text were rare in abstracts before 2020 but increased in recent years, especially after the release of ChatGPT. Some journal types use these tools more than others. These findings help raise awareness about the growing role of artificial intelligence in scientific communication.

## Introduction

The release of large language models (LLMs) such as (OpenAI, San Francisco, CA), Gemini (Google, Mountain View, CA), and Claude (Anthropic, San Francisco, CA), has brought a new era in natural language processing (NLP) and artificial intelligence (AI). These generative AI systems can produce human-like text with fluency, coherence, and contextual awareness, and have rapidly gained traction across a wide range of domains, including scientific research[1]. In the biomedical sciences, LLMs are increasingly being used to assist with literature reviews, manuscript drafting, and even peer review, raising both opportunities and challenges for the academic community[2, 3].

There has been significant controversy in the scientific world about the rising popularity of LLMs. A rising number of journals, including the Journal of Urology and European Urology now require disclosure of the use of LLMs when submitting abstracts. Concerns relating to factual inaccuracies (“ hallucinations”) from LLM use have been increasingly mediatized in the public. Furthermore, the true impact of ChatGPT on scientific writings has not been well defined at this point, thus raising concerns in its growing use [2]. However, some proponents of LLMs argue that its use can increase accessibility of scientific communication, particularly for non-native English speakers and inexperienced researchers [4].

Multiple studies have shown a growing use in LLMs in scientific writing; however, no data currently exists regarding its adoption within specific medical specialties such as urology [1, 2]. To address this gap, we conducted a longitudinal analysis of urology abstracts published between 2010 and 2024, employing a mixture model approach to estimate the proportion of AI-like text over time. Our study provides the first quantitative assessment of the rise of generative AI in urology literature.

## Methods

We conducted a retrospective analysis of urology abstracts published between 2010 and 2024 to assess the prevalence of AI-like text, inspired by the methodology described by Liang et al.[5]. The Web of Science platform from Clarivate was accessed in July 2025, and a search of the category “ Urology and Nephrology” was initiated. All abstracts pertaining to the domain of Urology, with a Clarivate (Clarivate Analytics, London, UK) 2024 Journal Impact Factor >= 2 were selected. Thirty-eight journals were selected and then grouped into 4 categories: General, Functional Urology, Men’s Health, Oncology, Endourology. All included journals can be found in Supplementary Table 1. PubMed abstracts were extracted from all journals using the Entrez API. The search strategy included English-language abstracts with full publication dates and was filtered to exclude non-research article types (e.g., letters, comments, errata).

**Table 1.**
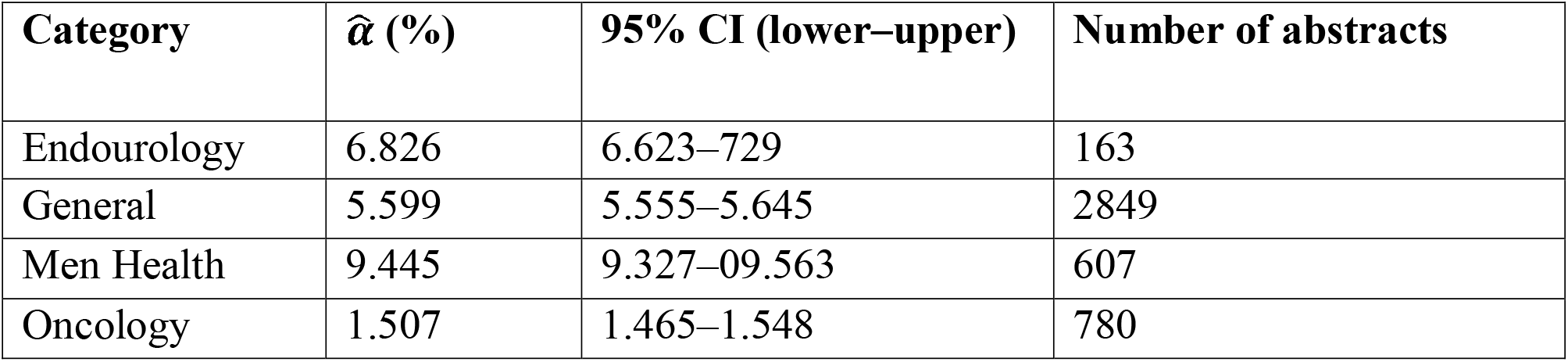
Estimated α by Journal Category, 2024.

**Table 2.** Estimated α by Journal Impact Factor Quartile, 2024.

| **IF Quartile (range)** | **$\hat{\alpha}$ (%)** | **95% CI (lower–upper)** | **Number of abstracts** |
| --- | --- | --- | --- |
| Q1 (2.0 – 2.31) | 5.931 | 0.05864–0.05997 | 1251 |
| Q2 (2.4 – 2.7) | 4.980 | 0.04910–0.05049 | 998 |
| Q3 (2.8 – 4.4) | 3.315 | 0.03253–0.03377 | 888 |
| Q4 (4.5 – 25.2) | 6.572 | 0.06501–0.06643 | 1310 |

### Data Preprocessing

Abstracts were preprocessed and standardized using (NLP) techniques, including removal of special characters, normalization of whitespace, and removal of duplicate entries based on PMID identifiers.

### Synthetic Reference Corpus Generation

To establish a reference distribution for AI-like text, we generated 10,000 synthetic abstracts using GPT-3.5-turbo-1106 (OpenAI, San Francisco, CA) with temperature settings (a parameter controlling output randomness) of 0.7, 0.8, and 0.9 to capture variations in AI outputs [6]. This model was selected because it represents the most popular, freely available early version of ChatGPT, and is likely the version that researchers relied on during the study period. The synthetic generation process used only paper titles as prompts to avoid contamination from existing abstracts. Each synthetic abstract was generated using the following prompt: “ Write a scientific abstract (200–250 words) for the urology paper titled *‘title’*.” The synthetic corpus was limited to titles from pre-2020 publications to ensure temporal separation from the analysis period, given that early LLMs started to become popular in 2020.

### Tokenization and Alpha Estimation Methodology

Text tokenization was performed using cl100k_base encoding from the tiktoken library which is the tokenizer used by GPT-3.5-turbo-1106. [7].

The proportion of AI-like text *(α*) was estimated using maximum likelihood estimation of a mixture model, as described by Liang et al. [5]. For each time period, the likelihood function was:

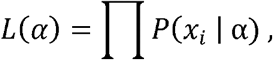

where the maximum likelihood estimate 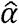 was obtained as:

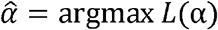

and where:

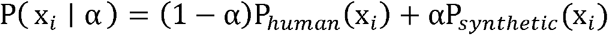

Here, P_*human*_ (x_*i*_)and P_*synthetic*_ (x_*i*_) represent the frequency of token x_*i*_ in the pre-2020 human corpus, and the synthetic reference corpus, respectively, while 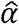 is the estimated proportion of AI-generated text. Laplace smoothing (with a constant of 0.5) was then applied.

### Lexical Diversity

To assess trends in lexical diversity over time, we calculated the type-token ratio (TTR) for urology abstracts in each year from 2010 to 2024. The TTR is defined as the number of unique word types divided by the total number of tokens, providing a simple measure of vocabulary richness. For each year, all abstracts were concatenated and tokenized using the tiktoken library, and the TTR was computed as the ratio of unique tokens to total tokens.

### Statistical Analysis

Alpha estimates 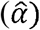 were calculated annually from 2010-2024 for overall trends. Confidence intervals were computed using the Wald formula:

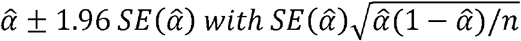

where n represents the total number of tokens in the analyzed corpus. Analyses stratified by IF and Journal category were performed for the year 2024. All statistical analyses were performed using Python 3.13 with scipy and pandas libraries.

### Validation of Alpha Estimation Methodology

To evaluate calibration, we constructed controlled mixtures of real and synthetic abstracts at known proportions (*α* = 0.00 to 1.00 in increments of 0.05) and estimated 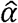 for each mixture. The alpha estimator was applied to the real abstracts from 2006–2009 to assess baseline specificity. The validation set included 1,000 synthetic abstracts generated using GPT-3.5-turbo-1106 with pre-2010 titles and the same prompt structure as the main analysis.

### Ethical statement

This study was conducted using only publicly available data from published urology abstracts and synthetic text generated by large language models. No human subjects, patient data, or confidential information were involved. As such, institutional review board (IRB) approval and informed consent were not required.

## Results

### Study Cohort and Corpora

The initial Entrez query retrieved 98,903 abstracts. After excluding entries with empty text fields and duplicate PMIDs, 64,444 unique abstracts remained. No records were removed following the integration of impact factor and journal category data. These abstracts were distributed across journal impact factor quartiles (Supplementary Table 3).

To construct the human-written reference corpus (H), we compiled 42,495 urology abstracts published before 2020, encompassing approximately 14.9 million tokens and 38,175 unique word types. The synthetic reference corpus (S) consisted of 10,000 abstracts generated using GPT-3.5-turbo, totaling over 3.1 million tokens and 17,697 unique word types.

### Token frequency distribution

Analysis of token frequency distributions revealed distinct patterns across human-written, synthetic, and post-ChatGPT urology abstracts (**Figure 1**). The post-ChatGPT abstracts, when visualized alongside these reference distributions, shows a bell curve intermediate between the human and synthetic corpora.

**Figure 1.**
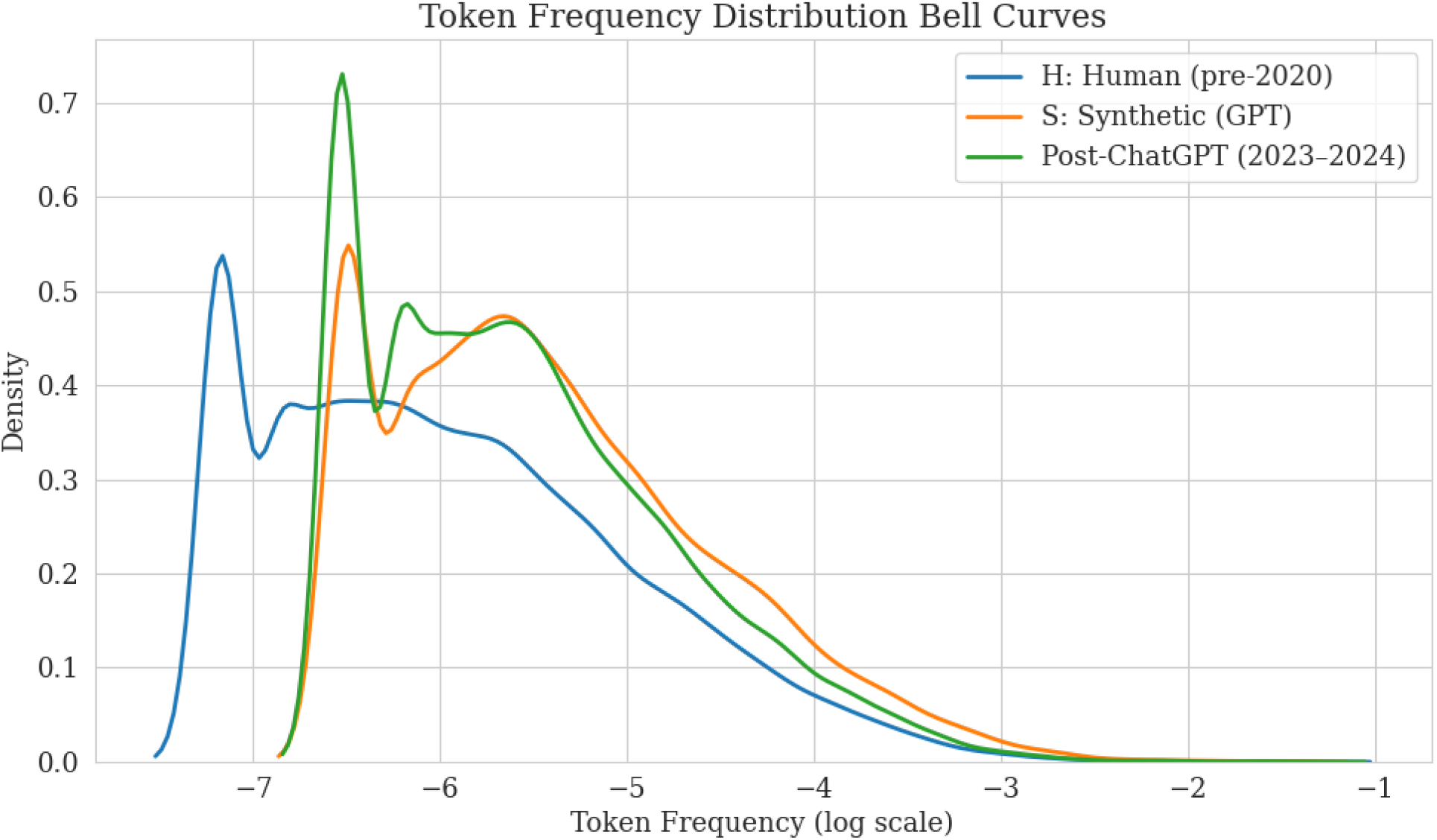
Bell curves of token frequency distributions for human-written, synthetic, and post-ChatGPT urology abstracts (log scale).

### Overall Trend of AI-like Text in Urology Abstracts

The estimated proportion of AI-like text 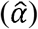 in urology abstracts remained negligible from 2010 to 2019, with 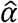 values consistently near zero. An inflection point occurred in 2020 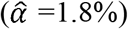, leading to a steady increase in the prevalence of AI-like language. This trend accelerated significantly after the public launch of ChatGPT in late 2022. The mean estimated 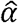 rose from 3.1% in 2022 to 3.7% in 2023, reaching a peak of 5.3% in 2024 (**Figure 2**).

**Figure 2.**
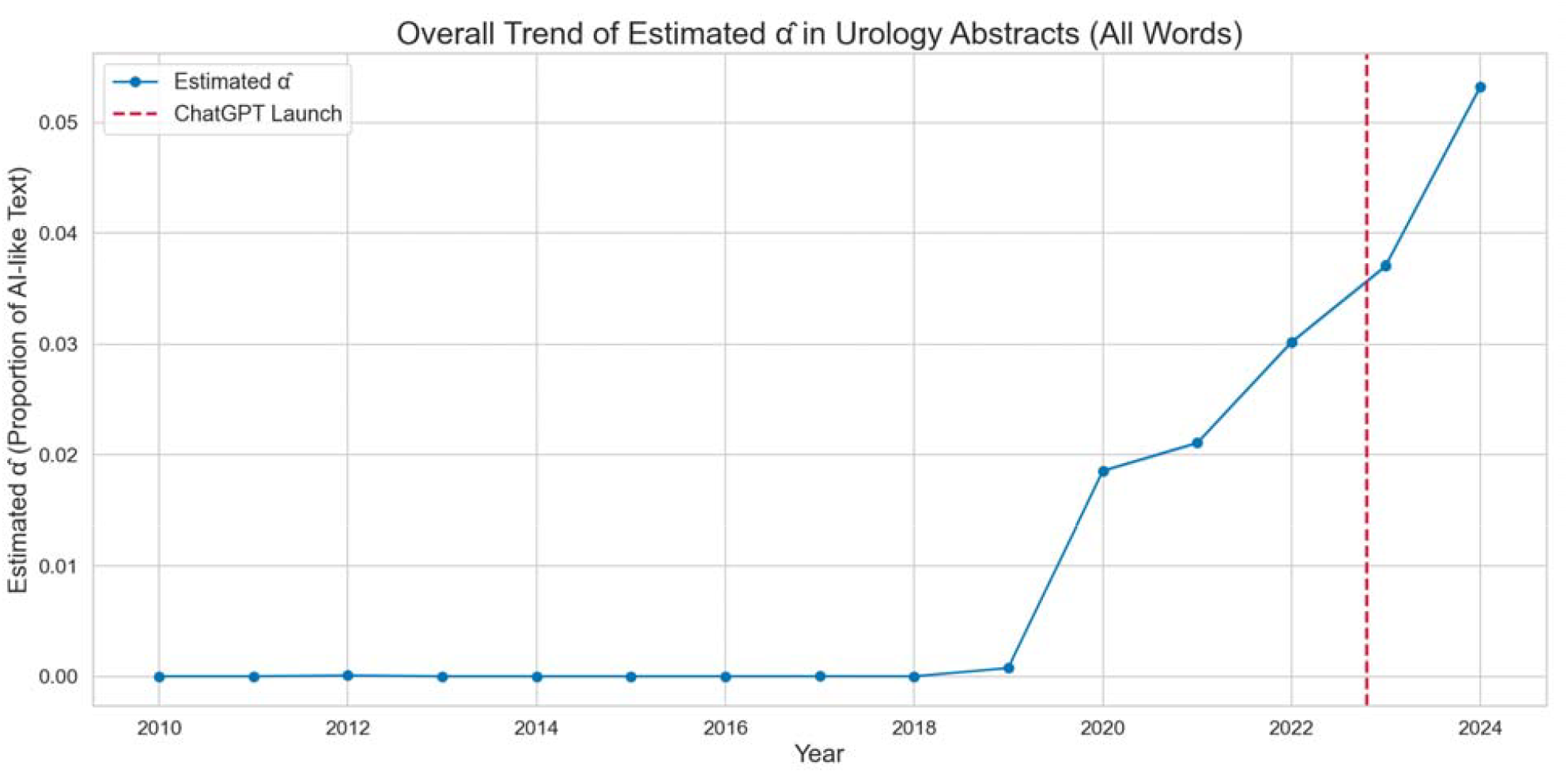
Overall trend of the estimated proportion of AI-like text (in urology abstracts from 2010 to 2024. The dashed red line indicates the launch of ChatGPT.

### AI-like Text by Journal Category and Impact Factor Quartile

The Functional Urology category was excluded from this subgroup analysis, as only a single journal in that domain was represented in the dataset. In 2024, the proportion of AI-like text varied considerably across urological subspecialties. Men’s Health journals had the highest estimated 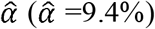, followed by Endourology 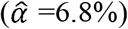 and General 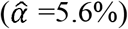. Oncology Urology journals reported the lowest levels (1.4%). When grouping the journals by quartiles, the bottom (Q1) and top (Q4) quartiles showed the highest prevalence of AI-like content (5.9% and 6.6%, respectively), whereas those in the middle quartiles (Q2 and Q3) had lower values (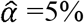 and 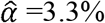).

### Lexical Diversity

In the pre-2020 human abstracts (H), a total of 2,285,951 adjective tokens representing 18,955 unique types and 318,290 adverb tokens representing 4,436 unique types were identified. In contrast, the synthetic GPT-3.5 corpus (S) contained 506,089 adjective tokens with 8,114 unique types and 63,135 adverb tokens with 1,233 unique types. Analysis of lexical diversity, as measured by the type-token ratio (TTR), demonstrated that the vocabulary richness of urology abstracts has remained relatively stable over the past 15 years (Supplementary Table 4). The TTR was 0.0163 in 2010 and fluctuated only modestly throughout the study period, with values ranging from 0.0139 to 0.0163. Although minor year-to-year variations were observed, there was no evidence of a sustained upward or downward trend, even in the years following the introduction of ChatGPT.

### Validation

In the negative control validation using 12,549 urology abstracts from 2006–2009, the estimated proportion of AI-generated text 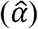 was 0.0060 (95% CI: 0.0060–0.0061), indicating a negligible false positive rate for the estimator in pre-LLM data. In the calibration experiment, where controlled mixtures of real and synthetic abstracts were created at known proportions, the estimated α□ closely tracked the true mixture proportion across the full range (Supplementary Figure 1). The area under the calibration curve was 0.533, suggesting that the estimator is well-calibrated and accurately estimates the true proportion of AI-generated text in mixed samples.

## Discussion section

This study provides the first quantitative evidence of a rapid and substantial increase in the prevalence of AI-like text in urology abstracts, coinciding with the public release of ChatGPT in late 2022. For nearly a decade, the proportion of AI-like text in urology abstracts remained near zero based on our estimations. Beginning in 2020, a modest increase was detected, but it was not until after the public release of ChatGPT in late 2022 that the prevalence of AI-like text accelerated in a significant manner. The early inflection point observed around 2020 may, in part, be attributable to the release and increasing accessibility of earlier LLMs such as OpenAI’s GPT-2 (released in 2019) and GPT-3 (released in 2020) [6]. While these models were not as widely adopted as ChatGPT, their availability may have enabled a small number of researchers to experiment with AI-assisted writing prior to the mainstream adoption seen with ChatGPT.

Our findings are consistent with but also extend, recent work in other biomedical fields. For example, Liang et al. employed a similar mixture model approach to estimate the prevalence of LLM-like text in PubMed abstracts, using a reference corpus of pre-2020 human-written abstracts and a synthetic corpus generated by prompting GPT-3.5-turbo with titles [5]. Their study, which analyzed over 100,000 abstracts across multiple specialties, reported a comparable post-2022 surge in AI-like text, with *α* values rising from near zero to over 17.5% in computer science by 2023. The Nature portfolio showed the among the least increase in α, with 6.3%. Kobak et al. introduced an alternative method to estimate LLM usage by analyzing excess vocabulary words that increased disproportionately after the release of ChatGPT across 15 million PubMed abstracts from 2010 to 2024 [1]. Their approach, which avoids reliance on ground-truth synthetic corpora, estimated that at least 13.5% of 2024 abstracts were LLM-assisted, with usage reaching over 40% in some subfields such as computational biology. Other recent investigations have attempted to use alternative approaches to detect AI-generated content in scientific writing. Gao et al. (2023) asked blinded reviewers to distinguish real abstracts from those generated by ChatGPT. While 68% of synthetic abstracts were correctly identified, 14% of genuine ones were misclassified as AI-generated, demonstrating the difficulty in reliably detecting LLM-generated text.[8]. These studies show that detection at the individual document level is difficult, and that our mixture modeling approach offers a complementary, population-level perspective, quantifying the aggregate impact of generative AI on the scientific literature. Our study builds on this foundation by providing a focused analysis within urology, incorporating stratification by journal category and impact factor, and extending the time series through 2024.

### Subgroup Analysis

On subgroup analysis, significant heterogenicity exists in terms of prevalence of LLM uptake. The Men’s Health category showed the highest proportion of AI-like text 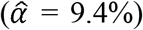, which could be explained by a higher proportion of journals being international-based (4/7, 57%). Authors less comfortable with English could derive significant benefit from the use of LLMs, as non-native speakers can face most difficulties in writing the Introduction and Discussion section as per a survey by John Flowerdew [9]. A study by Ziang Xu of 168 AI usage declarations across 8,859 Elsevier articles found that non-native English speakers and international author teams were significantly more likely to use generative AI tools [10].

On the other hand, Oncology-focused Urology journals had the lowest 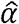 (1.4%), possibly due to involvement of large multidisciplinary teams, as well as having most of the leading oncology research centers in the United States, a high-income and predominantly English-speaking country. Stricter editorial oversight could also play a factor, however the effectiveness and impact of Editorial policy on the use of LLMs was not studied yet. These differences show that LLM integration is not uniform across different fields of urology and might be shaped by domain-specific cultural practices and linguistic demands. In terms of impact factor subanalyses, we see a U-shaped relationship between the IF quartile and 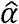. Journals in Q1 and Q4 showed higher levels when compared to journals in Q2 and Q3. Higher rated journals might attract authors who might be early adopters of the technology and use LLMs to accommodate higher research outputs. Lower IF journals having a high level of AI-like text could suggest lower editorial oversight regarding LLM use, as well as author’s decreased access to resources leading reliance of LLMs [11].

### Implications in the field of Urology

With the rise of AI in academic writing, significant opportunity arose, particularly for researchers who face barriers related to language proficiency or writing skills. As mentioned earlier in the discussion, non-native speakers can face significant barriers in disseminating research. In a cohort study by Li et al., a cohort of 25 third year medical students were asked to write a mini paper with and without the assistance of ChatGPT. The use of LLM was associated with higher scoring of the paper, as well as 92% of the students noting that the resource improved the quality of their writing, and 84% reporting advancements in their own language skills[4]. This resource could contribute to leveling the playing field of the global scientific community, which has historically been mostly English based. It is estimated that around 90% of natural sciences indexed publications are written in English[12]. A large-scale analysis by Lepp et al. examined over 80,000 peer reviews at top computer science conferences and found that submissions from authors in non–English-speaking countries were more likely to receive lower ratings and more critiques on clarity of the text [13].It is clear that the use of LLMs can serve as powerful aids in scientific writing, however the fast uptake we have seen in multiple fields brings significant liabilities. The most mediatized phenomenon around LLM use has been “ hallucinated content”, which consists of content generated by the model that appears plausible but is factually incorrect, fabricated, or unsupported by any real-world source. A study by Chelli et al. used a comparative analysis to test ChatGPT and Bard’s ability to replicate 11 systematic reviews on rotator cuff pathology, and found the models were unreliable, with high hallucination rates (28.6% to 91.4%) [14]. These high hallucination rates are very concerning as they could add a significant amount of false information in the scientific literature. Furthermore, the traditional way a manuscript generally has been written reflected the intellectual and cultural background of the authors, thus contributing to a rich and diverse scientific discourse. If generative AI becomes a default tool in manuscript generation, we might see a convergence towards a more limited vocabulary and lose nuances in our scientific language. Finally, as the boundary between human and machine-generated content becomes increasingly blurred, questions arise about who will be truly responsible for the ideas and arguments presented in scientific publications.

Several limitations of our study should be acknowledged. First, while the mixture model provides a robust estimate of the prevalence of AI-like text at the population level, it cannot attribute authorship or distinguish between direct and indirect AI assistance. Second, our reference corpora, though carefully constructed, may not capture the full diversity of writing styles or the evolving capabilities of different or more recent LLMs. Third, our analysis is limited to abstracts; the prevalence and impact of AI-generated text in full manuscripts, peer reviews, or other scientific communications may differ. Finally, the observational nature of our study precludes causal inference regarding the drivers of AI adoption.

## Conclusion

In conclusion, this study provides the first quantitative evidence of a rapid and substantial increase in the prevalence of textual patterns similar to AI-generated text in urology abstracts, with a marked inflection point following the public release of ChatGPT in late 2022. By applying a robust mixture modeling approach to a large, longitudinal dataset, we demonstrate that the integration of generative AI into scientific writing is not only widespread but accelerating, with significant variation across journal categories and impact factor quartiles. As generative AI becomes increasingly integrated into scientific writing, the urology community must ensure that advances in communication are matched by continued commitment to authorship integrity and scientific rigor.

## Data Availability

All data produced in the present study are available upon reasonable request to the authors

