## Supplemental File for "The Silent Author in Urology: Quantifying Large Language Model Influence at the Corpus Level"

Supplementary table 1:

Table 1. Lists of Journals included in the study

| **Journal Name** | **ISSN** | **eISSN** | **IF** | **Category** |
| --- | --- | --- | --- | --- |
| European Urology | 0302-2838 | 1873-7560 | 25.2 | General |
| Nature Reviews Urology | 1759-4812 | 1759-4820 | 14.6 | General |
| European Urology Oncology | NA | 2588-9311 | 9.3 | Oncology |
| Journal of Urology | 0022-5347 | 1527-3792 | 6.8 | General |
| Prostate Cancer and Prostatic Diseases | 1365-7852 | 1476-5608 | 5.8 | Oncology |
| European Urology Focus | NA | 2405-4569 | 5.6 | General |
| European Urology Open Science | 2666-1691 | 2666-1683 | 4.5 | General |
| International Braz J Urol | 1677-5538 | 1677-6119 | 4.5 | General |
| BJU International | 1464-4096 | 1464-410X | 4.4 | General |
| Minerva Urology and Nephrology | 2724-6051 | 2724-6442 | 4.2 | General |
| World Journal of Men's Health | 2287-4208 | 2287-4690 | 4.1 | Men Health |
| Therapeutic Advances in Urology | 1756-2872 | 1756-2880 | 3.5 | General |
| Sexual Medicine Reviews | 2050-0513 | 2050-0521 | 3.4 | Men Health |
| Journal of Sexual Medicine | 1743-6095 | 1743-6109 | 3.3 | Men Health |
| World Journal of Urology | 0724-4983 | 1433-8726 | 2.9 | General |
| Urologic Clinics of North America | 0094-0143 | 1558-318X | 2.9 | General |
| Current Urology Reports | 1527-2737 | 1534-6285 | 2.9 | General |
| Journal of Endourology | 0892-7790 | 1557-900X | 2.8 | Endourology |
| Asian Journal of Andrology | 1008-682X | 1745-7262 | 2.7 | Men Health |
| Clinical Genitourinary Cancer | 1558-7673 | 1938-0682 | 2.7 | Oncology |
| Research and Reports in Urology | 2253-2447 | 2253-2447 | 2.7 | General |
| Prostate International | 2287-8882 | 2287-903X | 2.6 | General |
| Aging Male | 1368-5538 | 1473-0790 | 2.6 | General |
| Prostate | 0270-4137 | 1097-0045 | 2.5 | General |
| International Journal of Impotence Research | 0955-9930 | 1476-5489 | 2.5 | Men Health |
| Asian Journal of Urology | 2214-3882 | 2214-3890 | 2.4 | General |
| Urologic Oncology - Seminars and Original Investigations | 1078-1439 | 1873-2496 | 2.3 | Oncology |
| Advances in Urology | 1687-6369 | 1687-6377 | 2.3 | General |
| Current Sexual Health Reports | 1548-3584 | 1548-3592 | 2.3 | Men Health |
| Urolithiasis | 2194-7228 | 2194-7236 | 2.2 | Endourology |
| International Journal of Urology | 0919-8172 | 1442-2042 | 2.2 | General |
| Current Opinion in Urology | 0963-0643 | 1473-6586 | 2.2 | General |
| International Neurourology Journal | 2093-4777 | 2093-6931 | 2.1 | Functional Urology |
| Investigative and Clinical Urology | 2466-0493 | 2466-054X | 2.1 | General |
| Scandinavian Journal of Urology | 2168-1805 | 2168-1813 | 2.1 | General |
| Sexual Medicine | N/A | 2050-1161 | 2.0 | Men Health |
| Urology | 0090-4295 | 1527-9995 | 2.0 | General |
| CUAJ-Canadian Urological Association Journal | 1911-6470 | 1920-1214 | 2.0 | General |

**Supplementary table 2:**

Number of abstracts per year per category

| Year | Functional Urology | Endourology | General | Men Health | Oncology |
| --- | --- | --- | --- | --- | --- |
| 2010 | 43 | 312 | 2772 | 522 | 154 |
| 2011 | 36 | 287 | 2922 | 526 | 189 |
| 2012 | 34 | 271 | 2890 | 521 | 225 |
| 2013 | 32 | 321 | 3014 | 553 | 393 |
| 2014 | 32 | 306 | 2977 | 566 | 437 |
| 2015 | 37 | 297 | 2933 | 567 | 418 |
| 2016 | 69 | 286 | 2795 | 442 | 397 |
| 2017 | 49 | 279 | 2713 | 347 | 567 |
| 2018 | 52 | 247 | 2672 | 329 | 503 |
| 2019 | 51 | 115 | 2651 | 378 | 535 |
| 2020 | 53 | 64 | 2893 | 454 | 560 |
| 2021 | 53 | 86 | 3270 | 508 | 635 |
| 2022 | 55 | 122 | 2766 | 439 | 595 |
| 2023 | 44 | 101 | 2644 | 430 | 541 |
| 2024 | 48 | 155 | 2673 | 492 | 704 |
| 2025 | 0 | 0 | 377 | 89 | 128 |

**Supplementary Table 3:**

Lexical diversity (TTR) year over year:

| Year | TTR |
| --- | --- |
| 2010 | 0.0163 |
| 2011 | 0.0155 |
| 2012 | 0.0143 |
| 2013 | 0.0140 |
| 2014 | 0.0144 |
| 2015 | 0.0145 |
| 2016 | 0.0151 |
| 2017 | 0.0152 |
| 2018 | 0.0155 |
| 2019 | 0.0158 |
| 2020 | 0.0154 |
| 2021 | 0.0139 |
| 2022 | 0.0149 |
| 2023 | 0.0152 |
| 2024 | 0.0140 |

Supplementary Figure 1:

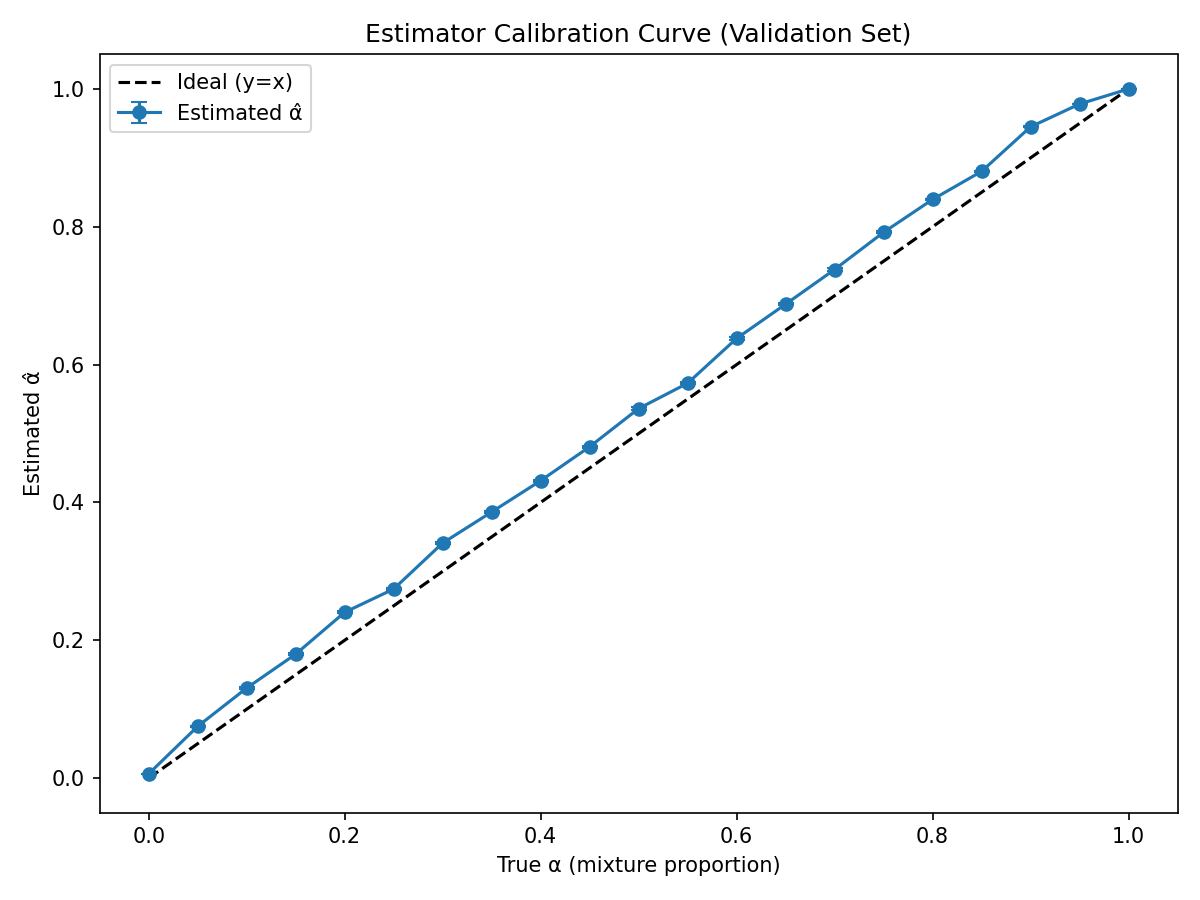

Figure 1. Calibration curve of the alpha estimator on the validation set (2006–2009).
